# Low *IGFBP7* expression identifies a subset of breast cancers with favorable prognosis and sensitivity to IGF-1 receptor targeting with ganitumab: Data from I-SPY2 and SCAN-B

**DOI:** 10.1101/2023.12.18.23300129

**Authors:** Christopher Godina, Michael N Pollak, Helena Jernström

## Abstract

There has been a long-standing interest in targeting the insulin-like growth factor-1 receptor (IGF-1R) signaling system in breast cancer due to its key role in neoplastic proliferation and survival. However, no IGF-1R targeting agent has shown substantial clinical benefit in controlled trials, and no treatment predictive biomarkers for IGF-1R targeting agents exist. IGFBP7 is an atypical insulin-like growth factor binding protein as it has a higher affinity for the IGF-1R than IGF ligands. We report that low *IGFBP7* gene expression identifies a subset of breast cancers for which the addition of ganitumab (an anti-IGF-1R monoclonal antibody) to chemotherapy substantially improved the pathological complete response rate compared to neoadjuvant chemotherapy alone. Furthermore, high *IGFBP7* expression predicted increased distant metastasis risk. If our findings are confirmed, decisions to halt the development of IGF-1 targeting drugs, which were based on disappointing results of prior trials that did not use predictive biomarkers, should be reviewed.

## Introduction

A significant portion of breast cancer patients relapse despite receiving optimal treatment adjuvant or neoadjuvant according to current guidelines ^1^. This fact highlights the need for new treatments and new biomarkers to better guide patient selection.

Dysregulation of insulin-like growth factor-1 receptor (IGF-1R) signaling has long been correlated with breast cancer growth, proliferation, and survival ^2,3^. Many agents have been developed to target IGF-1R ^4^. To date, no studies have demonstrated a substantial clinical benefit of adding IGF-1R targeting agents to existing treatments ^4^. A possible explanation is that response to chemotherapy is enhanced by IGF-IR targeting only in a subset of tumors, following the well-known precedent that adding HER2 targeting agents to chemotherapy is only helpful in HER2-positive tumors ^5–7^. However, no predictive biomarkers for IGF-IR targeting agents have been identified.

Recently, the Investigation of Serial Studies to Predict Your Therapeutic Response With Imaging And moLecular Analysis 2 (I-SPY2) trial showed a small increase in pathological complete response (pCR) rate when the anti IGF-1R monoclonal antibody ganitumab (AMG479) (given with metformin to reduced treatment-induced hyperglycemia) was added to chemotherapy, but the effect size failed to reach the prespecified threshold for a positive result ^8^. Further, eleven putative IGF-1R signaling axis biomarkers were tested, but none were predictive of ganitumab benefit ^8^.

I-SPY2 has been able to identify treatment predictive biomarkers for emerging treatments due to its unique design^9–12^, and to evaluate efficacy of new treatments^13–16^. To further this goal, the I-SPY2-990 Data Resource has been made publicly available^13^.

It has previously been shown^17^that unlike other IGFBPs, IGFBP7 can bind to the IGF-1R-, but the *in vivo* consequences of the IGFBP7 - IGF-1R interaction are unclear. There are also gaps in knowledge concerning complex interactions between IGFBP7 and IGF-1R in the presence of anti-IGF-1R antibodies such as Ganitumab. IGFBP7 binds insulin-like growth factor-1 (IGF-1) and the IGF-1R in a mutually exclusive manner^17^. Compared to other IGFBPs, IGFBP7 binds insulin with higher affinity than IGFs ^17,18^. The binding of IGFBP7 decreases activation and internalization of IGF-1R in response to IGF-1/2 but at the same time sensitizes IGF-1R to insulin stimulation ^17,18^. IGFBP7 was shown to prolong the surface retention of the IGF-1R under insulin/IGF1 stimulation resulting in prolonged IGF-1R signaling in leukemia ^18^. It also has been shown that IGFBP7 promotes the persistence of IGF-1R at the cell surface, prolonging insulin/IGF stimulation, and enhancing Akt activation leading to mitogenic and pro-survival effects ^18,19^.

Although we have presented prior evidence that high IGFBP7 expression by breast cancer tissue or high circulating levels of this protein are related to poor prognosis ^20,21^, this protein has not previously been studied in relation to efficacy of IGF-1R targeted therapies. We previously reported that the prognostic value of circulating IGFBP7 was only seen for patients who had cancers with positive IGF-1R membrane status ^21^, which suggested that the interplay between IGFBP7 and IGF-1R merits further investigation. We hypothesized that IGFBP7 might compete with IGF-1R monoclonal antibody binding to IGF-1R, decreasing its efficacy and at the same time promoting tumor growth and metastatic potential.

## Materials and methods

### I-SPY2

The I-SPY2 trial (NCT01042379) is an open-label, multicenter, adaptively randomized phase 2 trial of neoadjuvant therapy, evaluating multiple investigational arms in parallel. Neoadjuvant paclitaxel followed by doxorubicin/cyclophosphamide (with Trastuzumab in HER2+ disease) serves as the common control arm ^15,16^. Investigational agent(s) are combined with this regimen ^15,16^. The primary endpoint is pCR (ypT0/is, ypN0) ^15,16^. Eligibility for I-SPY2 includes age 18 years or older, stage II or III breast cancer and primary tumor size >2.5 cm by clinical examination or >2.0 cm by imaging ^15,16^Patients with hormone receptor (HR)+ cancers are eligible only if they have a poor prognosis estimated by the MammaPrint (MP) 70-gene based prognostic signature^13^. Subsequent adjuvant endocrine treatment was at the discretion of the treating physician. Ganitumab was tested in combination with metformin since ganitumab can induce hyperglycemia ^8^. In the I-SPY2 study of the IGF-IR blocking antibody ganitumab, patients with HER2+ tumors were not included since there is no safety data concerning the combination of ganitumab and trastuzumab ^8^. Gene expression profiling of core needle biopsies taken from the primary tumor before treatment was performed using Agilent 44K expression arrays ^13^. Transcriptomic and clinical data of patients enrolled in the I-SPY2 trial were obtained from the Gene Expression Omnibus (GEO) database (GSE194040). For the analysis reported here, we studied patients from the ganitumab/metformin plus chemotherapy arm and the chemotherapy-alone control arm, excluding patients with HER2+ disease (Figure 1).

**Figure 1.**
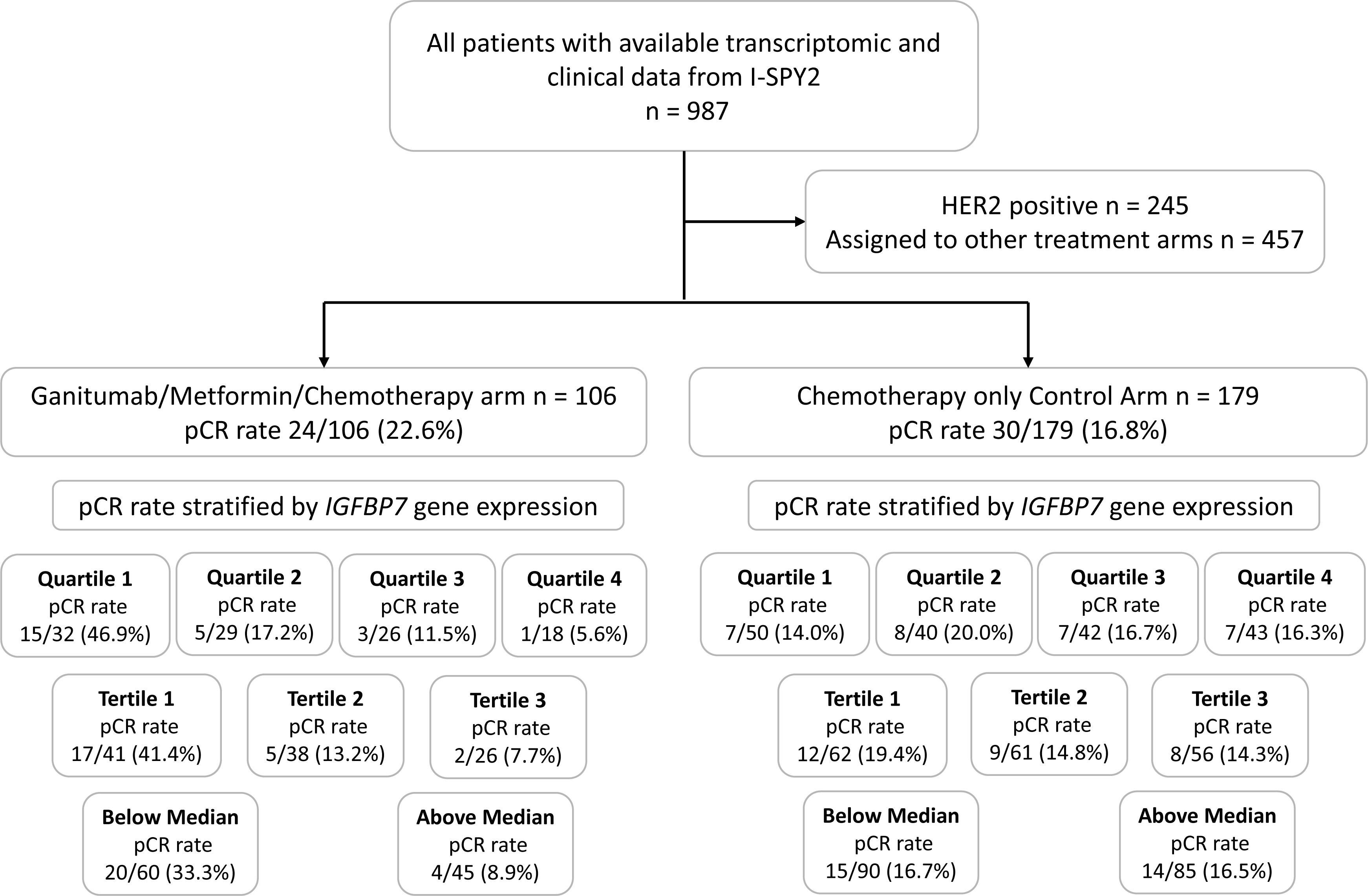
Flowchart of included and excluded patients in I-SPY2. pCR rates in the ganitumab/metformin plus chemotherapy arm and chemotherapy control arm, according to *IGFBP7* expression categories.

### SCAN-B

The Swedish Cancerome Analysis Network – Breast (SCAN-B; NCT02306096) is an ongoing population-based cohort. SCAN-B prospectively includes breast cancer patients diagnosed and treated at nine Swedish hospitals ^22,23^. All newly diagnosed breast cancer patients are invited to participate ^23^. Tumor specimens or core needle biopsies (in case of neoadjuvant treatment) from the patients’ tumors are obtained in conjunction with routine clinical sampling ^22,23^. The samples are subject to gene expression profiling using RNA-seq according to custom SCAN-B workflow ^22–24^. Gene expression levels were expressed in fragments per kilobase of exon per million mapped reads (FPKM) ^24^. Clinical data was collected from the Swedish National Quality Registry for Breast Cancer ^22–24^. Curated RNA-seq and clinical data were accessed from Staaf *et al*. ^24^. A subset of 5326 patients with available follow-up for distant metastasis, no bilateral cancer, and gene expression profiles (GEXs) from a primary invasive breast cancer was analyzed (Supplementary Figure 1). If multiple GEXs for a single patient were available, the GEX with the highest RNA concentration was chosen ^24^, leaving one GEX per patient for analysis.

### Data analysis

In both I-SPY2 and SCAN-B, eight gene expression modules representing different biological functions in breast cancer were calculated as previously described ^25^. The correlations between *IGFBP7* gene expression, gene expression of 15 other proteins involved in the IGF/Insulin pathway, and the eight modules were calculated using Pearson’s correlation.

In the regression analyses, *IGFBP7* gene expression was modelled both as a continuous variable and categorized as quartiles (quartile 1 (Q1), quartile 2 (Q2), quartile 3 (Q3), and quartile 4 (Q4)) to allow for non-linear effects. The lowest expression of *IGFBP7* (Q1) was used as reference. Quartiles were created separately for I-SPY2 and SCAN-B based on the entire datasets.

Crude and adjusted odds ratios (OR) with 95% confidence intervals (CI) for pCR in I-SPY2 was estimated using logistic regression. The multivariable models were adjusted *a priori* for potential confounders either previously known factors associated with likelihood of achieving pCR and/or variables that somewhat differed by *IGFBP7* quartiles: HR+, HER2+, trial arm (control as reference), PAM50 subtype (Luminal A as reference), MP ultra-high risk (MP2), ISPY-2 immune (Immune+), and DNA repair deficiency (DRD+) signatures ^13^. To investigate if there was any interaction between *IGFBP7* gene expression and efficacy of ganitumab/metformin plus chemotherapy treatment in achieving pCR, interaction analyses were performed in the ganitumab/metformin plus chemotherapy arm and the HER2 negative subset of the control arm. The interaction was tested in both crude and adjusted models by including an interaction term and comparing the models with or without the interaction term using the Likelihood ratio (LR) test. The *P*-values for the interaction term and LR test are reported. Due to sparse data, PAM50 subtype could not be included in the interaction analyses.

The endpoints used for survival analyses in SCAN-B were recurrence-free interval (RFI), distant metastasis-free interval (DMFI), and overall survival (OS) ^24^. Cox proportional hazards regression was used to estimate crude and adjusted Hazard ratios (HRs) with 95% CI. The multivariable models were adjusted for *a priori* selected standard clinicopathological factors: age (binned in 5-year intervals), tumor characteristics; lymph node status (pN1/2/3), tumor size (pT2/3/4), Grade (III), estrogen receptor (ER)+, progesterone receptor (PR)+, HER2+, PAM50 subtype (Luminal A as reference), PAM50 ROR category (High vs Low/Intermediate); and (neo)adjuvant treatments; endocrine treatment, chemotherapy, and trastuzumab.

Differential gene expression (DGE) analysis was performed using the ‘Limma-Voom’ package ^26^. The criteria used to define differentially expressed genes (DEGs) between the QT4 and Q1 of *IGFBP7* expression was false discovery rate (FDR) of[≤[0.05 and log2 fold change (log2FC)[≥[1.5 for up-regulated genes and log2FC[≤ −1.5 for down-regulated genes. The ‘clusterprofiler’ ^27^ package was used to perform gene set enrichment analysis (GSEA). Gene sets were grouped according to Hallmark Signatures ^28^. The results were visualized using ‘EnhancedVolcanoplot’, ‘pHeatmap’ and ‘clusterprofiler’ ^27^ packages. *In silico* profiling of different carcinoma ecosystems (including estimates of relative abundance) were derived from gene expression profiles of tumors from I-SPY2 using a deconvolution-based method, ECOTYPER (with standard parameters) ^29^. ECOTYPER applies a machine learning framework for large-scale identification of cell states and cellular ecosystems from bulk gene expression data ^29^.

All data analyses were conducted in R version 4.2.2. *P*-values <0.05 was considered statistically significant. All *P*-values were two-tailed. This study followed the Reporting Recommendations for Tumor Marker Prognostic Studies (REMARK) criteria ^30^.

## Results

Figure 1 summarizes the pCR rates in patient subsets across the I-SPY2 trial. In all patients (across all treatment arms), *IGFBP7* (continuous and in quartiles) expression was not associated with odds of achieving pCR in the multivariable model (Supplementary Table 1). Descriptive statistics for clinicopathological factors are presented in Table 1, Supplementary Table 2 for I-SPY2. There was an interaction between *IGFBP7* expression (continuous and in quartiles) and efficacy of ganitumab/metformin plus chemotherapy treatment in achieving pCR in both the crude (LR *P*=0.011 for both continuous and quartiles) and multivariable models (LR *P*=0.017 for continuous and LR *P*=0.015 for quartiles) (Table 2). In the ganitumab/metformin plus chemotherapy arm 22.6% of patients achieved pCR compared to 16.8% in the chemotherapy alone control arm. However, higher *IGFBP7* expression conferred lower odds of achieving pCR in the ganitumab/metformin plus chemotherapy arm, adjusted OR 0.38 (95% CI 0.17–0.80) but not in the chemotherapy alone control arm, adjusted OR 1.23 (95% CI 0.63–2.45; *P*_interaction_=0.016) (Figures 1-2 and Table 2). Likewise, Q2-4 of *IGFBP7* expression compared to Q1 also conferred lower odds of achieving pCR in the ganitumab/metformin plus chemotherapy arm (Q4 adjusted OR 0.09 (95% CI 0.00–0.58)) but not in the chemotherapy-alone control arm (adjusted OR 1.38 (95% CI 0.41–4.73; *P*_interaction_=0.031)) The crude and adjusted ORs for Q2-3 and other clinicopathological factors are provided in Figure 1 and Table 2. When stratified by quartiles, in the ganitumab/metformin plus chemotherapy arm 46.9% of *IGFBP7* Q1 tumors achieved pCR compared to 17.2% of *IGFBP7* Q2 tumors, 11.5% of *IGFBP7* Q3 tumors, and only 5.6% of *IGFBP7* Q4 tumors (Figure 2B and Table 2). In the chemotherapy alone control arm, 14.0% *IGFBP7* Q1 tumors, 20.0% of *IGFBP7* Q2 tumors, 16.7% *IGFBP7* Q3 tumors, and 16.3% of *IGFBP7* Q4 tumors achieved pCR (Figure 1, Figure 2C and Table 2). When divided by breast cancer subtype (high risk HR+HER2-vs. TNBC), the ability of IGFBP7 expression to identify breast cancers more likely to respond to ganitumab/metformin plus chemotherapy than to chemotherapy alone, was more apparent in TNBC, where 66.7% of *IGFBP7* Q1 tumors and no *IGFBP7* Q4 tumors achieved pCR, but the numbers are too small to draw definitive conclusions (Table 3). In an exploratory analysis, stratified by *IGFBP7* expression quartiles, ganitumab/metformin plus chemotherapy treatment conferred higher odds of achieving pCR in tumors with low *IGFBP7* expression (Q1-2) (adjusted OR 2.64 (95% CI 1.16–5.49)) but not in in tumors with high *IGFBP7* expression (Q3-4), (adjusted OR 0.44 (95% CI 0.11–1.40)). The improved efficacy of ganitumab/metformin plus chemotherapy treatment compared to standard chemotherapy in achieving pCR was confined to the ≈ 25% of patients in the lowest quartile of IGFB7 expression, where the pCR rate was 46.9% in the ganitumab/metformin plus chemotherapy group, compared to 14% in the chemotherapy alone group (Figure 1 and Table 4).

**Figure 2.**
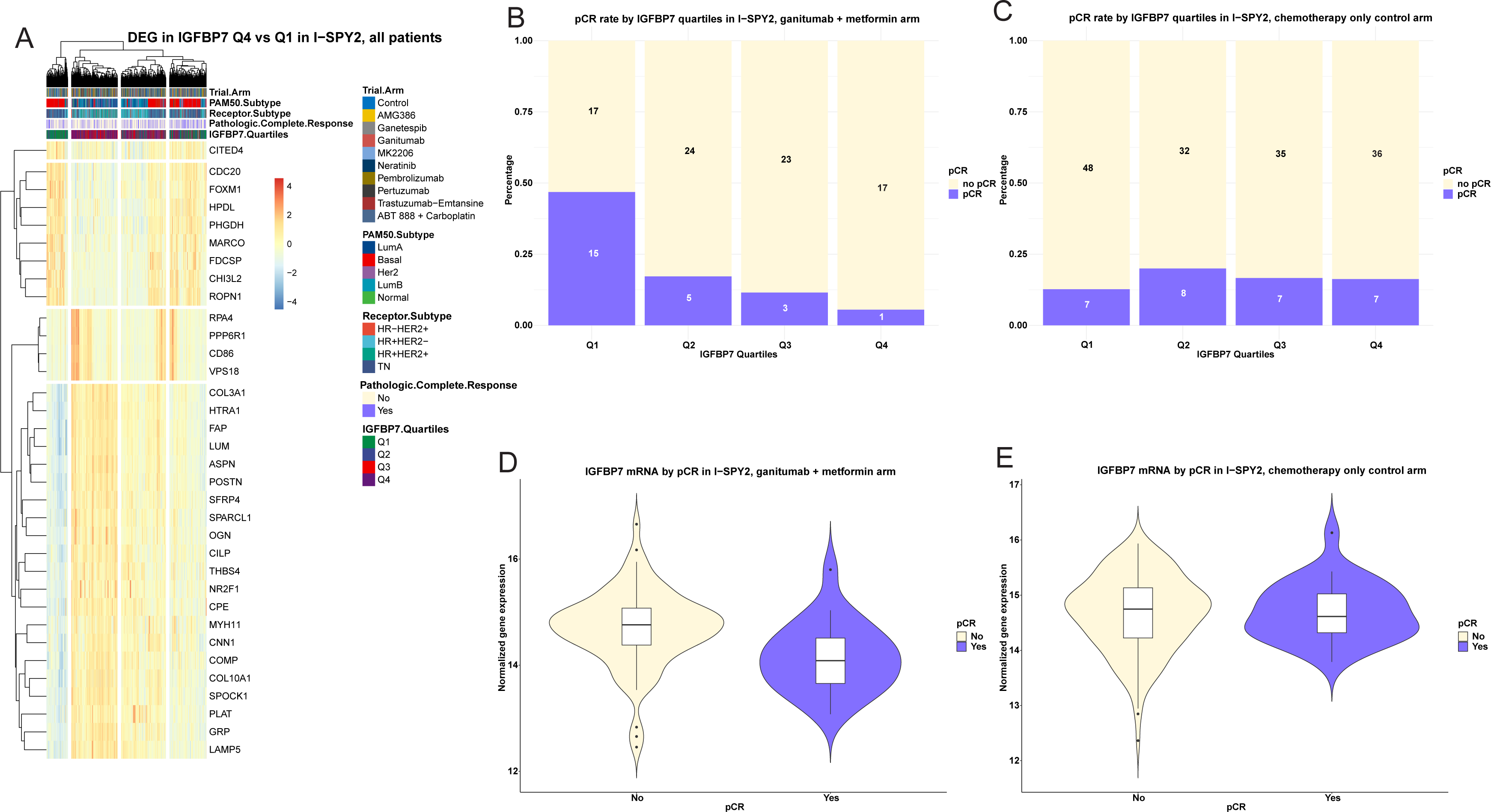
*IGFBP7* expression in relation to pCR Heatmap of differentially expressed genes (DEG) in *IGFBP7* Q4 compared to in *IGFBP7* Q1 tumors in (**A**) I-SPY2. Patients are along columns and biomarkers are along the rows. Red indicates higher expression and blue lower. Annotation tracks reflect pCR (purple), Receptor Subtype, PAM50 subtype, and trial arm (light red: ganitumab/metformin plus chemotherapy). *IGFBP7* expression (continuous) by pCR in (**B**) the ganitumab/metformin plus chemotherapy arm and in (**C**) the control arm. pCR rate by *IGFBP7* quartiles in (**D**) the ganitumab/metformin plus chemotherapy arm and in (**E**) the control arm.

**Table 1.**
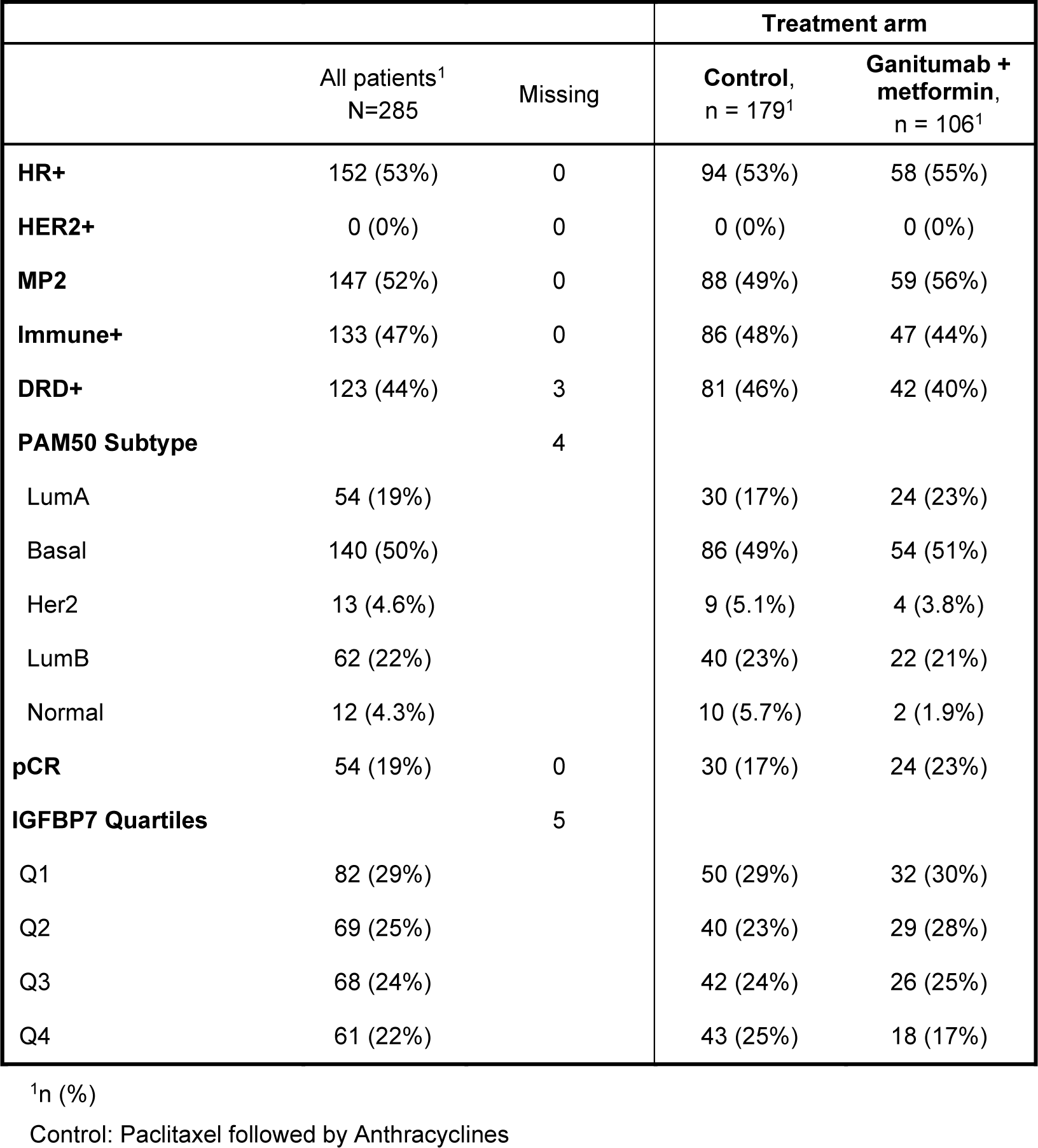
Descriptive statistics of chemotherapy only control arm and ganitumab plus metformin arm in relation to clinicopathological characteristics in I-SPY2.

**Table 2.**
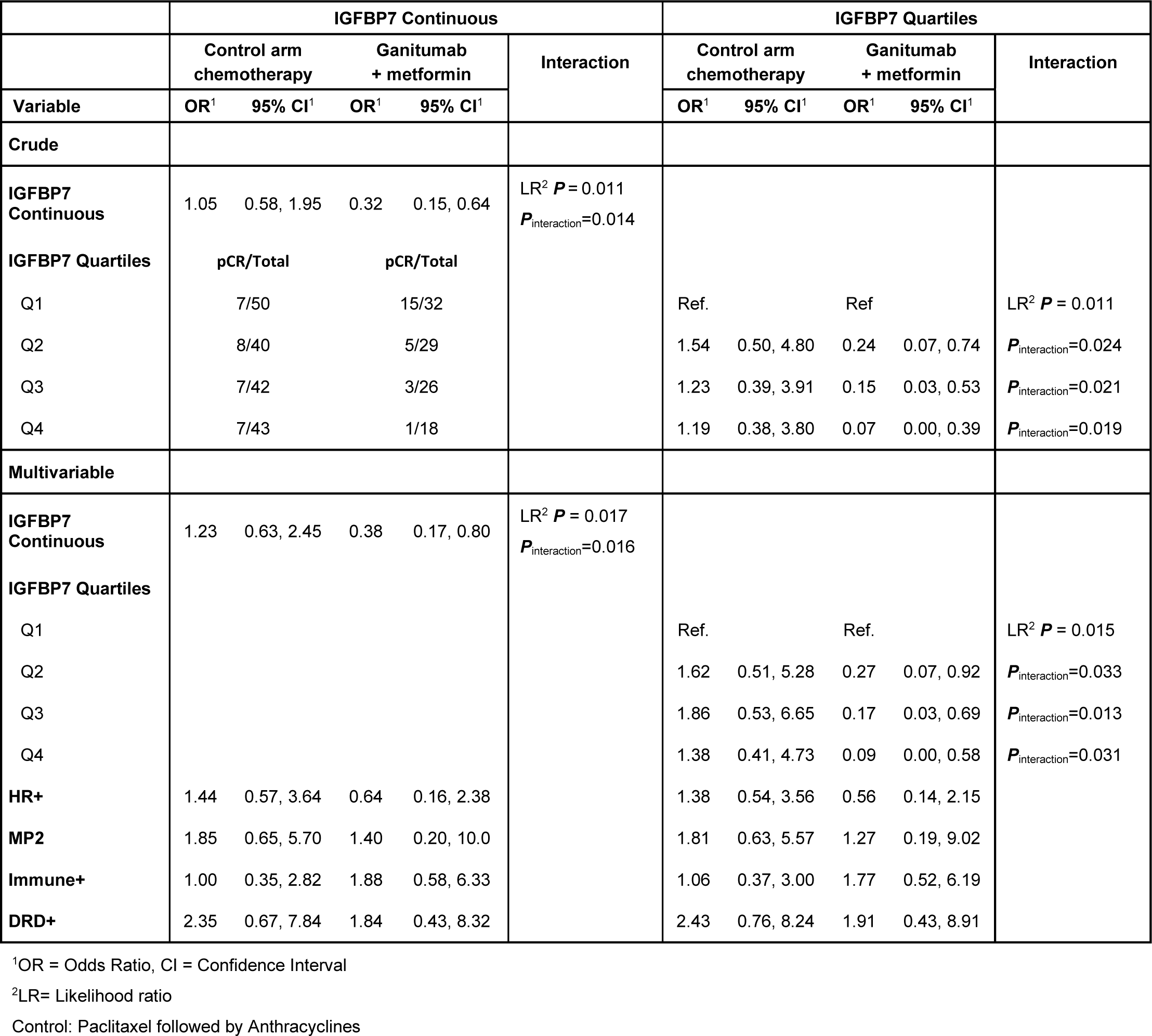
Odds ratio of achieving a pCR in relation IGFBP7 expression in control and ganitumub+metformin arms.

**Table 3.** pCR in relation *IGFBP7* expression in the chemotherapy only control arm and chemotherapy/ganitumub/metformin arm stratified by receptor subtype.

|  | HR+HER2– |  | TNBC |  |
| --- | --- | --- | --- | --- |
|  | Control arm | Ganitumab<br>+ metformin | Control arm | Ganitumab<br>+ metformin |
| ***IGFBP7* Quartiles** | **pCR/Total (%)** | **pCR/Total (%)** | **pCR/Total (%)** | **pCR/Total (%)** |
| Q1 | 5/21 (23.8%) | 3/14 (21.4%) | 2/29 (6.9%) | 12/18 (66.7%) |
| Q2 | 3/21 (14.3%) | 2/16 (12.5%) | 5/19 (26.1%) | 3/13 (23.1%) |
| Q3 | 2/31 (6.4%) | 2/15 (13.3%) | 5/11 (45.5%) | 1/10 (10%) |
| Q4 | 4/19 (21.0%) | 1/12 (8.3%) | 3/24 (12.5%) | 0/5 (0%) |

**Table 4.**
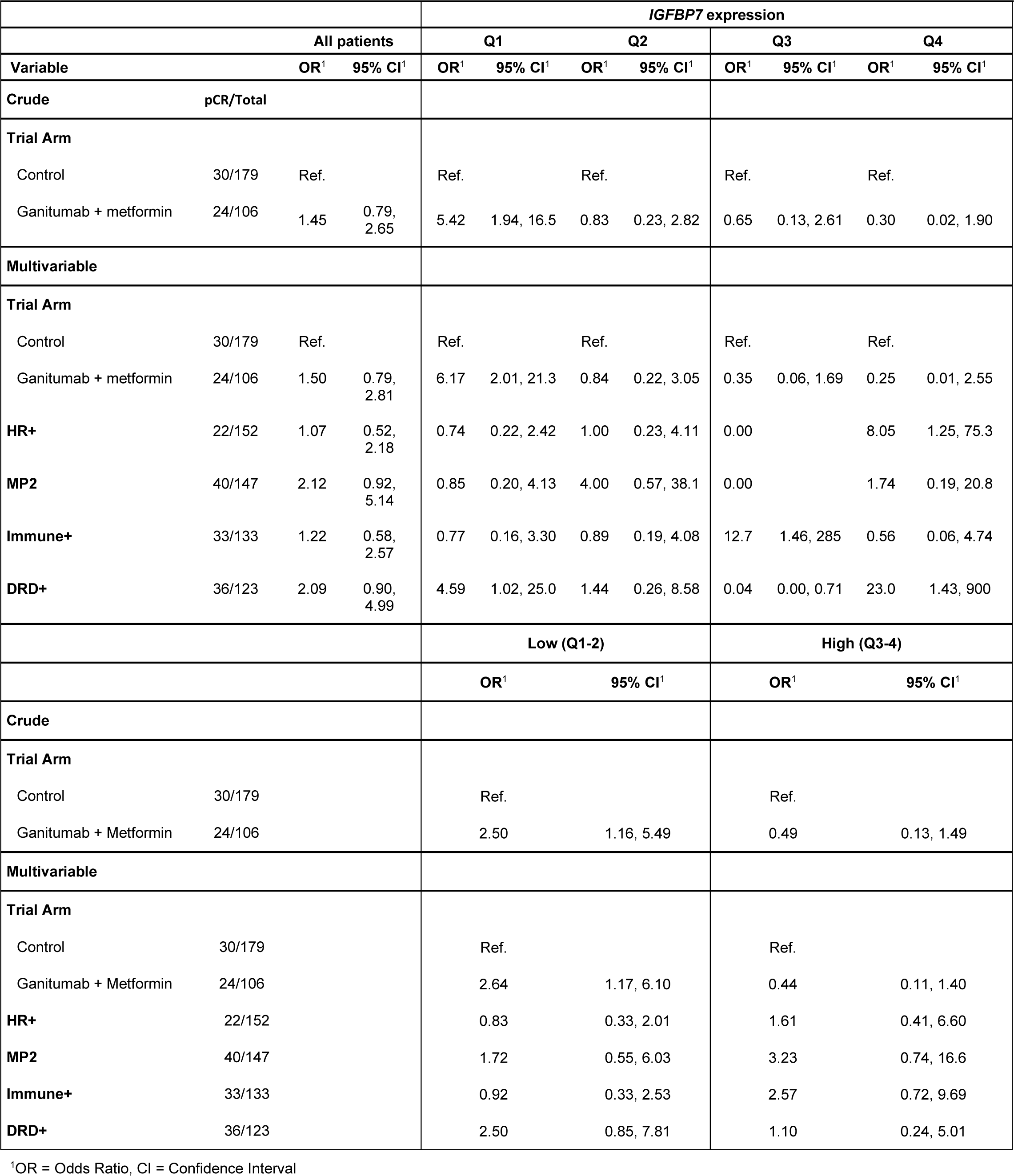
Odds ratio of achieving a pCR in the ganitumab+metformin arm compared to the chemotherapy only control arm, stratified by quartile and median of *IGFBP7* gene expression, I-SPY2.

In SCAN-B, the median follow-up for the 4158 patients still at risk was 5.45 (inter quartile range 5.07–8.15) years and descriptive statistics are presented in Supplementary Table 3 for SCAN-B. In the univariable survival analyses of *IGFBP7* expression in the SCAN-B cohort, *IGFBP7* expression Q4 compared to Q1 was not associated with increased risk of recurrence (HR 0.96 (95% CI 0.76–1.22)) or distant metastasis (HR 1.00 (95% CI 0.77–1.32)) (Supplementary Table 4). However, after adjustment for age, clinicopathological factors and treatment in the multivariable models, high *IGFBP7* expression (Q4 compared to Q1) was associated with significantly increased risk of recurrence (HR 1.37 (95% CI 1.04–1.82)) and distant metastasis (HR 1.60 (95% CI 1.15–2.22)) (Supplementary Table 4). When modeled as a continuous variable, *IGFBP7* expression showed similar associations with clinical outcome (Supplementary Table 5). Most importantly, *IGFBP7* (continuous) gene expression in the adjusted model also conferred a significantly increased risk of recurrence (HR 1.29 (95% CI 1.09–1.53)) and distant metastasis (HR 1.41 (95% CI 1.16–1.73)) (Supplementary Table 5).

Having demonstrated that low *IGFBP7* expression is predictive of a benefit of adding ganitumab to chemotherapy in neoadjuvant breast cancer treatment, we sought to determine if *IGFBP7* expression was related to other breast cancer characteristics. In both ISPY-2 and SCAN-B, *IGFBP7* expression was positively correlated with *IGFBP3-6* and *IGF1* and *IGF2* expression (all *r*_s_ ≥0.14) (Supplementary Figure 2 A-B). *IGFBP7* expression was highest in Normal-like followed by Luminal A subtype in both the ISPY-2 trial and SCAN-B cohort (Supplementary Figure 2 C-D). Likewise, the correlations between *IGFBP7* expression and the eight gene modules were similar in ISPY-2 and SCAN-B. *IGFBP7* expression was positively correlated with stroma, lipid, and early response to growth signaling modules and negatively correlated with mitotic checkpoint and progression modules (Supplementary Figure 2 E-F). The correlations suggest an association with aggressive tumor microenvironment (TME) and increased growth factor signaling activation. *IGFBP7* expression did not vary across HR-positive and HER2-positive subtypes (Supplementary Figure 2 G-H).

Differential gene expression (DGE) analyses were performed comparing *IGFBP7* Q4 versus Q1 tumors in I-SPY2 and SCAN-B. Notably, higher expression of several genes coding for proteins involved in endothelial cell regulation and extracellular matrix remodeling, *e.g*. *PDGFRB, COMP, FAP, COL1A1, OGN, LUM, COL3A1, SPOCK1, COL1A2, DCN, ANGPTL1,* and *SPON1* were seen in *IGFBP7* Q4 tumors, supporting a potential association with an active TME (Figure 2-3 and Supplementary Tables 6-7). Significantly enriched gene sets in *IGFBP7* Q4 tumors included EMT and TGF-β signaling (Figure 3 and Supplementary Tables 8-9). Tumor tissue composition was estimated by ECOTYPER^29^. High *IGFBP7* gene expression (continuous and in quartiles) was associated with dominance of Carcinoma Ecotype (CE) 6, CE 1, followed by CE 10 (P<0.001; Figure 4). This indicates that tumors with high *IGFBP7* gene expression have a microenvironment enriched for stromal cells while deficient in immune cells, characterized by transforming growth factor beta signaling (Figure 4). Conversely, tumors with low *IGFBP7* gene expression (continuous and in quartiles) were dominated by CE 2 and CE 9, indicating basal-like features and a pro-inflammatory response (Figure 4).

**Figure 3.**
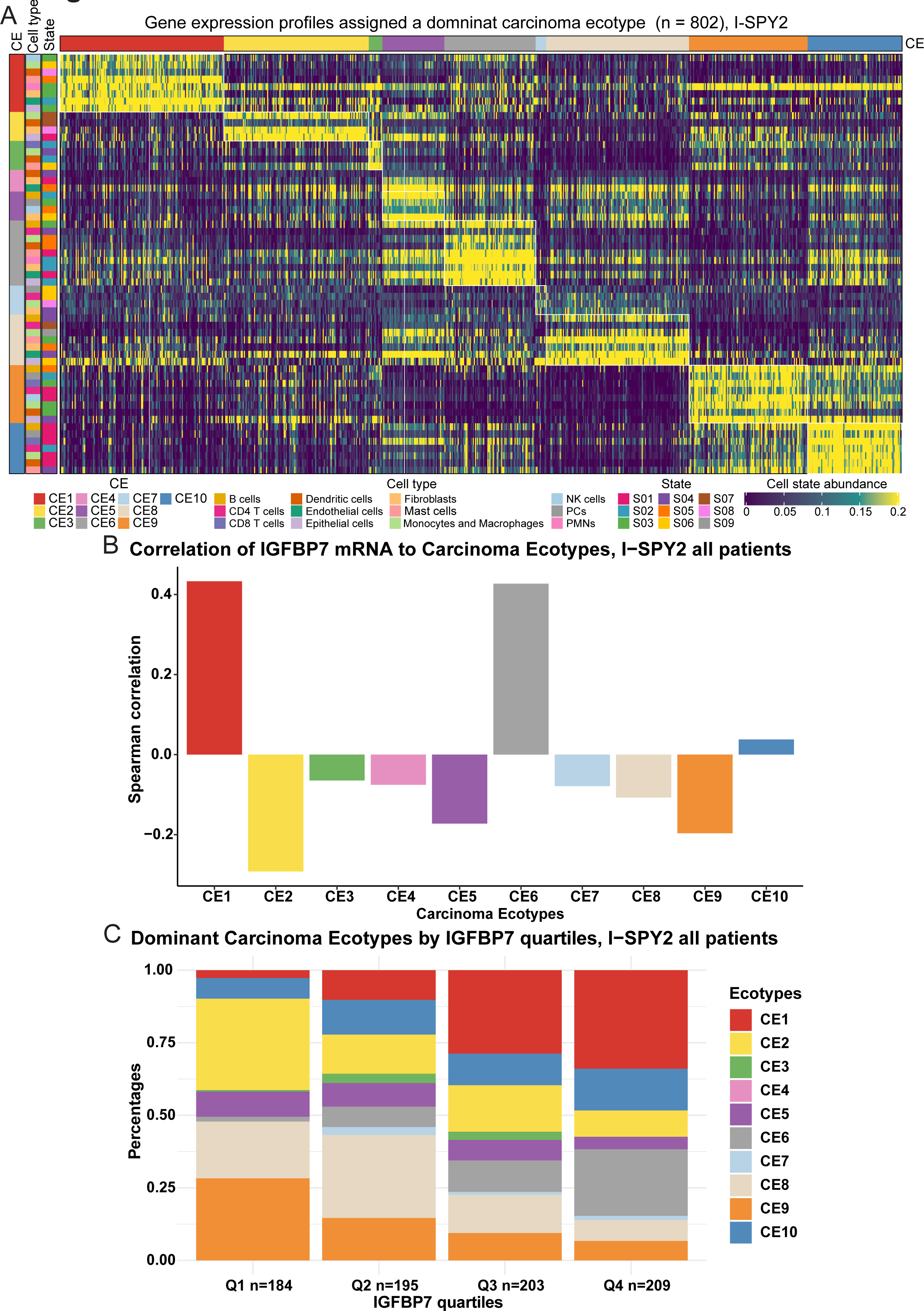
Molecular analyses of *IGFBP7* expression in I-SPY2 and SCAN-B Volcano plots showing significantly up- and downregulated genes in *IGFBP7* Q4 compared to in *IGFBP7* Q1 tumors in (A) I-SPY2, and (B) SCAN-B. Dot plots showing activated and suppressed Hallmark Signatures in *IGFBP7* Q4 compared to in *IGFBP7* Q1 tumors in (C) I-SPY2, and (D) SCAN-B. (D).

**Figure 4.**
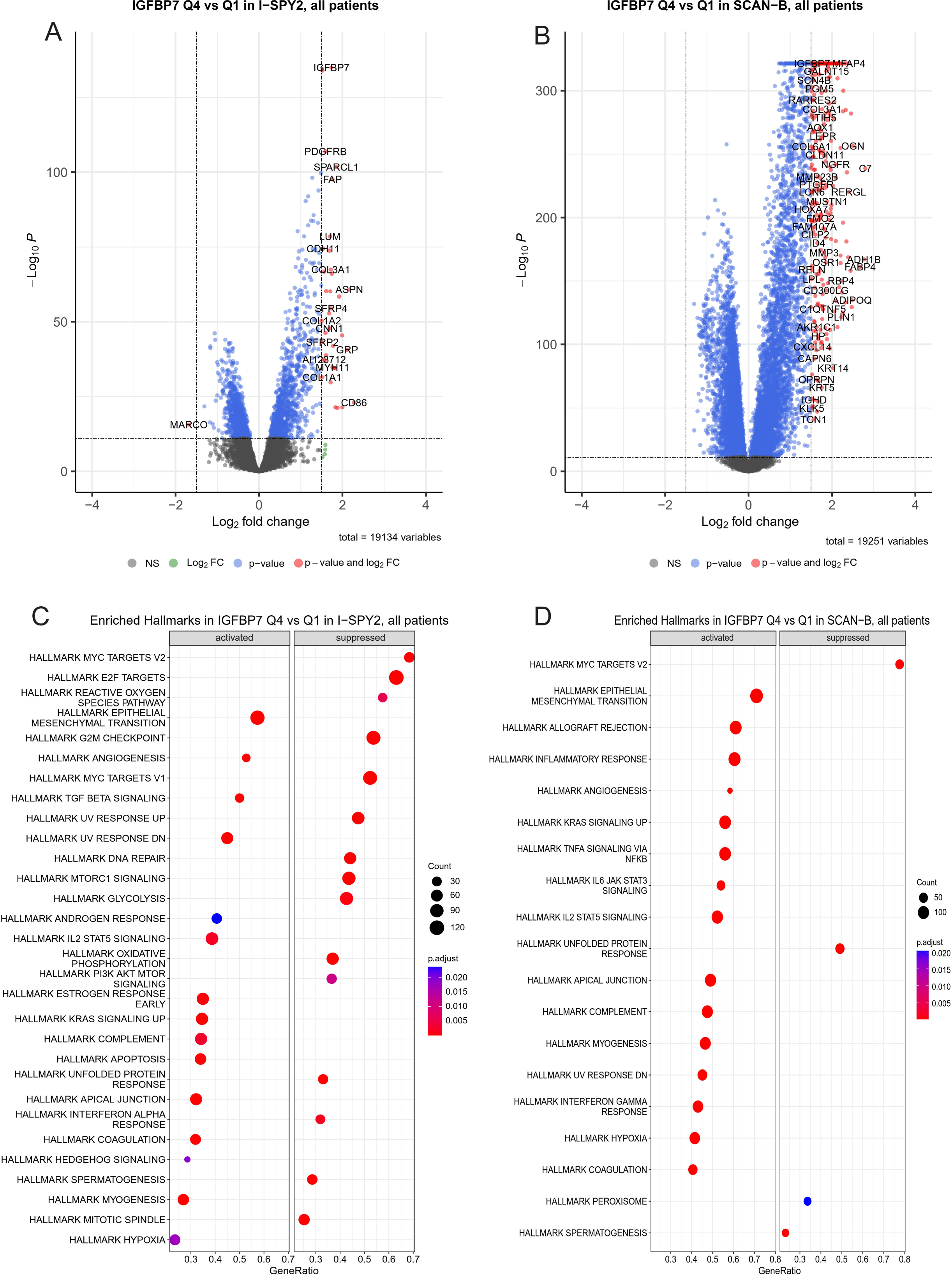
Tumor microenvironment composition in relation to *IGFBP7* gene expression Heatmap (A) of carcinoma ecotypes (CE), cell types, and cell states in patients (across all arms) in I-SPY2 whose tumors were assigned a dominant ecotype. Pearson correlations of IGFBP7 gene expression (continuous) and relative abundance of each CE. The dominant CE (C) in breast cancers by IGFBP7 quartiles.

## Discussion

We report, using data from the I-SPY trial, that breast cancer patients with tumors showing low *IGFBP7* gene expression had more than double the probability of achieving a pCR with neoadjuvant treatment using the combination of ganitumab, metformin, and chemotherapy than using neoadjuvant chemotherapy alone. In contrast, *IGFBP7* expression was unrelated to the probability of chemotherapy alone achieving a pCR. To our knowledge, this is the first study to date that identifies a specific biomarker related to efficacy of IGF-1R targeting treatment. By the use of *IGFBP7* as a predictor of ganitumab efficacy, we identified a substantial proportion of HER2 negative breast cancers including both poor prognosis HR^+^HER^-^ cancers (54.7% of patients) and TNBC (45.3% of patients) where the addition of ganitumab/metformin plus chemotherapy substantially improves pCR rates. Furthermore, we observed that high *IGFBP7* expression was associated with worse clinical outcome in SCAN-B, a large contemporary population-based breast cancer cohort.

Our finding that low *IGFBP7* expression defines a subset of patients where the addition of ganitumab to chemotherapy approximately doubles the pCR rate, shows the utility of the I-SPY2 trial design. Furthermore, our results demonstrate that detailed examination of the publicly available I-SPY2-990 Data Resource, either hypothesis-driven (as in this case), or agnostically, can provide opportunities to test hypotheses that were not proposed before or during the execution of the trial. The initial assessment of IGF signaling axis in the I-SPY2 ganitumab arm^8^ was negative but failed to include IGFBP7, a non-canonical IGFBP. Had *IGFBP7* been included in the initial assessment of predictive biomarkers in I-SPY2, it would have identified the distinct subgroup where ganitumab showed benefit. Our findings add to the list of important studies^11,13–16^ where the I-SPY2 trial detected activity of novel agents^10,11,15,16^, even if the benefit is shown to be confined to a biomarker-defined subset of patients.

The molecular features of tumors with high *IGFBP7* expression suggest an aggressive TME that facilitates metastasis. The ECOTYPER data suggest that strong evidence that IGFBP7 expression is a useful classifier distinct subtypes of the breast cancer microenvironment. This is consistent with previous studies showing that both systemic and tumor-specific IGFBP7 protein levels were poor prognostic markers in breast cancer^20,21^, and are also in line with our results from SCAN-B. Despite early studies^17–21^, there are many gaps in knowledge concerning the manner by which IGFBP7 modulates signaling by the IGF-1R family, and potentially the efficacy of anti-IGF-IR antibodies. Our findings provide a strong rationale for further experimental work to elucidate how IGFBP7 diminishes the effect of ganitumab and biological influence of IGFBP7 on the TME.

IGFBP7 merits further study as a treatment predictive biomarker in other cancer types such as colon, ovarian and prostate cancer, as well as sarcomas, where IGF-1R targeting agents have been investigated^4,31^. We emphasize that the lack of data concerning HER2-positive breast cancers does not imply lack of activity in these patients; they simply were not eligible for the ganitumab arm of the I-SPY2 study because of non-availability of clinical safety data regarding co-administration of ganitumab and HER2 targeting agents. In fact, prior literature^32^ has suggested benefit of co-targeting IGF-IR and HER2 signaling.

The strengths of this study include the use of contemporary data from both the I-SPY2 trial and SCAN-B cohort. SCAN-B allows for the evaluation of biomarkers in a contemporary real-world setting due to the large-scale RNAseq analysis of consecutively enrolled breast cancer patients ^22–24^. Further, I-SPY2 evaluates ganitumab in a randomized controlled setting, providing an ideal platform for investigation of treatment specific biomarkers for ganitumab. The *IGFBP7* expression showed stable associations with clinicopathological factors and molecular features across SCAN-B and ISPY-2.

The current study, while hypothesis driven, is a retrospective analysis of existing datasets and thus the results require further validation in prospective studies. Future work should include investigation of circulating IGFBP7 in relation to ganitumab response, as we did not have access to ISPY-2 blood samples. Another limitation is the lack of data on long-term outcome in I-SPY2, but it has been previously shown that pCR has a strong association with distant metastasis-free survival^33^. Due to the combination of ganitumab and metformin plus chemotherapy in the treatment arm, it is not possible to determine with certainty whether *IGFBP7* expression identifies tumors sensitive to ganitumab alone, metformin alone or the combination. However, the MA.32 randomized clinical trial did not show benefit of adjuvant metformin compared to standard therapy in early-stage breast cancer^34^, suggesting that the observed effect in I-SPY2 was driven by ganitumab.

There are few treatment options beyond chemotherapy in TNBC. Recently PARP inhibitors and immune checkpoint inhibitors have been incorporated in clinical practice^10,11,16,35–37^. Ganitumab plus chemotherapy combinations have a fairly benign side effect profile ^8^, and may improve outcomes for TNBC and other cancers, provided that IGFBP7 is validated as predictive biomarker. The main side effect is temporary hyperglycemia which can be managed with metformin.

In conclusion, low *IGFBP7* gene expression identifies a subset of breast cancers for whom the addition of ganitumab and metformin to neoadjuvant chemotherapy results in a significantly improved pCR rate compared to neoadjuvant chemotherapy alone. This justifies laboratory studies to address gaps in knowledge concerning roles of IGFBP7 in neoplasia, and the relevance of this protein to IGF-1R targeting agents. Such research, together with the results presented here, may lead to a review of decisions to halt the development of IGF-1 targeting drugs, which were based on disappointing results of prior trials that did not use predictive biomarkers.

## Data and code availability

Clinical and RNA-seq data from SCAN-B is accessible from Staaf *et al.* ^24^, available at Mendeley Data. The clinical and transcriptomic data from I-SPY2 are available from the GEO database (GSE194040). This paper does not report original code. Any additional information required to reanalyze the data reported in this paper is available upon request.

## Ethics statement

Ethical approvals for I-SPY2 and SCAN-B were obtained in relation to the primary projects and publications ^13,15,16,22–24^. No other separate approval was obtained for this specific study since it is based on previously published data. All participants signed written informed consent. The study was conducted in accordance with the ethical principles of the Declaration of Helsinki.

## Authors’ contributions

**C Godina**: Conceptualization, Data curation, Formal analysis, Methodology, Software, Resources, Visualization, Writing – original draft. **MN Pollak**: Conceptualization, Supervision, Writing – review & editing. **H Jernström**: Conceptualization, Funding acquisition, Formal analysis, Project administration, Supervision, Writing – original draft, Writing – review & editing. All authors have read and agreed to the published version of the manuscript.

## Data Availability

Clinical and RNA-seq data from SCAN-B is accessible from Staaf et al. 21, available at Mendeley Data. The clinical and transcriptomic data from I-SPY2 are available from the GEO database (GSE194040). This paper does not report original code. Any additional information required to reanalyze the data reported in this paper is available upon request. 21 Staaf, J., et al. RNA sequencing-based single sample predictors of molecular subtype and risk of recurrence for clinical assessment of early-stage breast cancer. NPJ Breast Cancer 8, 94 (2022).

https://www.ncbi.nlm.nih.gov/geo/query/acc.cgi?acc=GSE194040

https://data.mendeley.com/datasets/yzxtxn4nmd/3

## Acknowledgments

We thank the I-SPY2 consortium for generously making their data publicly available. We would also like to thank the I-SPY2 data and safety monitoring committee, trial coordinators, project oversight committee, and investigators, and all the patients who volunteered to participate in I-SPY2.Furthermore,we would like to acknowledge the patients who provided consent to participate in the SCAN-B study, the central SCAN-B laboratory at the Division of Oncology, Lund University for sample processing and RNA sequencing, the Swedish National Quality Register for Breast Cancer for clinical and histopathological data, Regional Cancer Center South, and the South Swedish Breast Cancer Group.

This study was funded by the Swedish Cancer Society (CAN 20 0763 and CAN 23 2952), the Faculty of Medicine at Lund University, the Mrs Berta Kamprad Foundation, and the South Swedish Health Care Region (Region Skåne ALF 40620). Dr. Pollak acknowledges funding from the Terry Fox Foundation. The funders had no role in study design and conduct of the study, data collection and analysis, data interpretation, or manuscript preparation and decision to submit the manuscript for publication.

## Abbreviations

DEGs: Differentially expressed genes
DGE: Differential gene expression
DMFI: Distant metastasis-free interval
ER: Estrogen receptor
EMT: Epithelial-mesenchymal transition
FC: Fold change
FDR: False discovery rate
FPKM: Fragments per kilobase of exon per million mapped reads
GEO: Gene Expression Omnibus
GEX: Gene expression profile
GSEA: Gene set enrichment analysis
HER2: Human epidermal growth factor receptor 2
HR: Hazard ratio
HR: Hormone receptor
IGF-1: Insulin-like growth factor-1 IGF-2 - Insulin-like growth factor-2
IGFBP7: Insulin-like growth factor receptor binding protein-7
IGF-1R: Insulin-like growth factor-1 receptor
I-SPY2: Investigation of Serial Studies to Predict Your Therapeutic Response With Imaging And moLecular Analysis 2
OR: Odds ratio
OS: Overall survival
pCR: pathological complete response
PR: Progesterone receptor
RFI: Recurrence-free interval
RNA-seq: Massive parallel paired-end sequencing of mRNA
SCAN-B: Swedish Cancerome Atlas Network - Breast
TME: Tumor microenvironment
TNBC: Triple-negative breast cancer
Q: Quartile

---

**Supplementary Figure 1.** Flowchart of included and excluded patients in SCAN-B

**Supplementary Figure 2.** *IGFBP7* expression in relation to molecular features Pearson correlations of *IGFBP7* gene expression (continuous) and 15 gene in the IGF/Insulin pathway (INS, INSR, IRS1, IRS2, IGF1, IGF2, IGFALS, IGF1R, IGF2R, IGFBP1, IGFBP2, IGFBP3, IGFBP4, IGFBP5, and IGFBP6) in (A) I-SPY2, and (B) SCAN-B. Pearson correlations of *IGFBP7* gene expression (continuous) and the eight gene modules (Stroma, Lipid, Immune Response, Mitotic Checkpoint, Mitotic Progression, Basal, Early Response, Steroid Response) in (C) I-SPY2, and (D) SCAN-B. *IGFBP7* expression (continuous) by PAM50 subtype in (E) I-SPY2 and (F) SCAN-B. SCAN-B. *IGFBP7* expression (continuous) by receptor subtype in (G) I-SPY2 and (H) SCAN-B.

