## Supplementary Table 1 for "Low *IGFBP7* expression identifies a subset of breast cancers with favorable prognosis and sensitivity to IGF-1 receptor targeting with ganitumab: Data from I-SPY2 and SCAN-B"

Supplementary Table S6-9 is available separately as an .xlsx file (Additional File 1).

### Supplementary figure legends

**Supplementary Figure 1.** Flowchart of included and excluded patients in SCAN-B

**Supplementary Figure 2.** *IGFBP7* expression in relation to molecular features

Pearson correlations of *IGFBP7* gene expression (continuous) and 15 gene in the IGF/Insulin pathway (INS, INSR, IRS1, IRS2, IGF1, IGF2, IGFALS, IGF1R, IGF2R, IGFBP1, IGFBP2, IGFBP3, IGFBP4, IGFBP5, and IGFBP6) in (**A**) I-SPY2, and (**B**) SCAN-B. Pearson correlations of *IGFBP7* gene expression (continuous) and the eight gene modules (Stroma, Lipid, Immune Response, Mitotic Checkpoint, Mitotic Progression, Basal, Early Response, Steroid Response) in (**C**) I-SPY2, and (**D**) SCAN-B. *IGFBP7* expression (continuous) by PAM50 subtype in (**E**) I-SPY2 and (**F**) SCAN-B. SCAN-B. *IGFBP7* expression (continuous) by receptor subtype in (**G**) I-SPY2 and (**H**) SCAN-B.

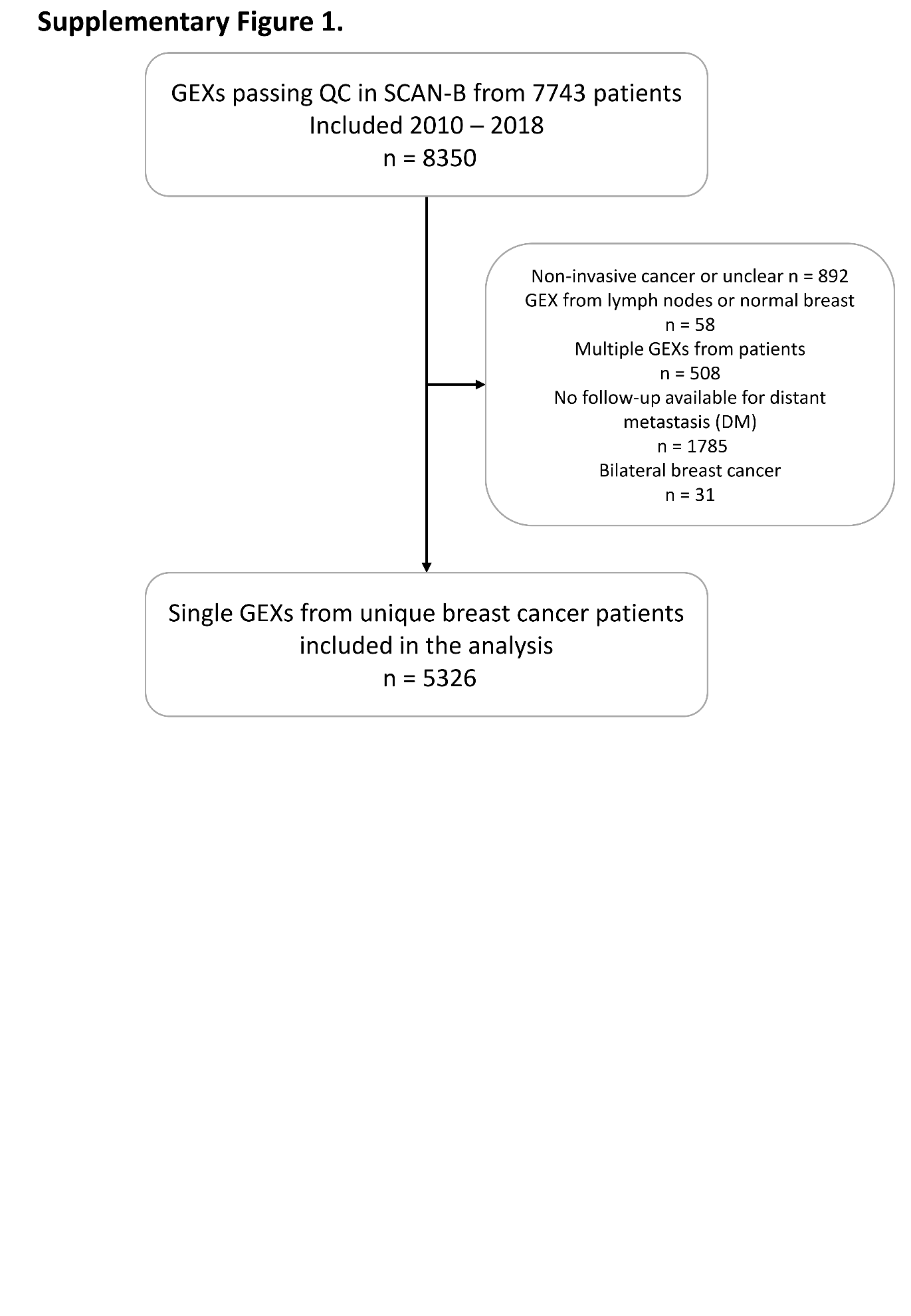

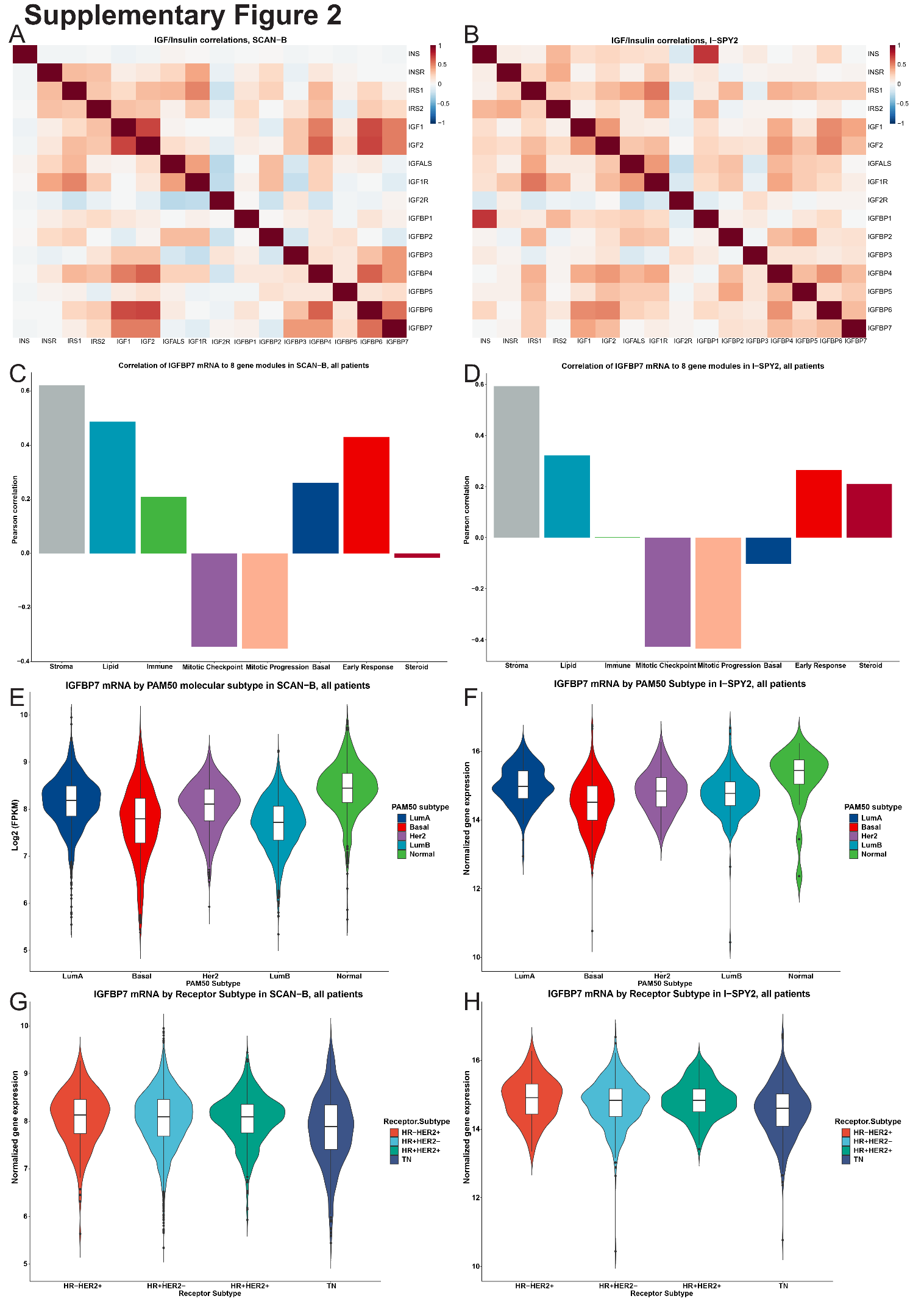

| **Supplementary Table 1** Odds ratio of achieving a pCR in relation *IGFBP7* gene expression, all patients (arms) in I-SPY2 | | | | |
| --- | --- | --- | --- | --- |
|  | **IGFBP7 Continuous** | | **IGFBP7 quartiles** | |
| **Variable** | **OR**^1^ | **95% CI**^1^ | **OR**^1^ | **95% CI**^1^ |
| **IGFBP7 Continuous** | 0.99 | 0.79, 1.25 |  |  |
| **IGFBP7 Quartiles** |  |  |  |  |
| Q1 |  |  | Ref. |  |
| Q2 |  |  | 0.77 | 0.50, 1.20 |
| Q3 |  |  | 1.19 | 0.76, 1.85 |
| Q4 |  |  | 0.96 | 0.60, 1.52 |
| **HR+** | 0.72 | 0.46, 1.10 | 0.71 | 0.46, 1.09 |
| **HER2+** | 3.32 | 1.88, 5.85 | 3.34 | 1.90, 5.90 |
| **MP2** | 1.54 | 0.97, 2.46 | 1.60 | 1.00, 2.56 |
| **Immune+** | 2.89 | 2.03, 4.15 | 2.88 | 2.02, 4.14 |
| **DRD+** | 1.64 | 1.03, 2.65 | 1.69 | 1.05, 2.73 |
| **PAM50 Subtype** |  |  |  |  |
| LumA | Ref. |  | Ref. |  |
| Basal | 1.86 | 0.83, 4.21 | 1.84 | 0.82, 4.17 |
| Her2 | 3.32 | 1.70, 6.60 | 3.34 | 1.71, 6.65 |
| LumB | 1.58 | 0.86, 2.95 | 1.65 | 0.90, 3.09 |
| Normal | 2.00 | 0.59, 6.43 | 2.02 | 0.60, 6.50 |
| **Trial Arm** |  |  |  |  |
| Control | Ref. |  | Ref. |  |
| AMG386 | 2.10 | 1.19, 3.73 | 2.15 | 1.22, 3.82 |
| Ganetespib | 1.74 | 0.91, 3.28 | 1.69 | 0.89, 3.20 |
| Ganitumab + metformin | 1.76 | 0.92, 3.32 | 1.75 | 0.92, 3.30 |
| MK2206 | 1.89 | 1.01, 3.54 | 1.85 | 0.99, 3.47 |
| Neratinib | 1.98 | 1.08, 3.64 | 2.01 | 1.10, 3.70 |
| Pembrolizumab | 5.38 | 2.70, 10.8 | 5.30 | 2.66, 10.7 |
| Pertuzumab | 4.91 | 2.09, 11.8 | 5.21 | 2.20, 12.6 |
| Trastuzumab-emtansine | 3.82 | 1.65, 8.99 | 3.94 | 1.69, 9.32 |
| Carboplatin + ABT888 | 3.55 | 1.80, 7.04 | 3.56 | 1.80, 7.06 |
| ^1^OR = Odds Ratio, CI = Confidence Interval  Control: Paclitaxel (+Trastuzumab if HER2+) followed by doxorubicin and cyclophosphamide | | | | |

| **Supplementary Table 2.** Descriptive statistics of IGFBP7 quartiles in relation to clinicopathological characteristics in I-SPY2 | | | | | | |
| --- | --- | --- | --- | --- | --- | --- |
|  |  |  | **IGFBP7 Quartiles** | | | |
|  | All patients^1^ | Missing | **Q1**,  n = 244^1^ | **Q2**,  n = 244^1^ | **Q3**,  n = 243^1^ | **Q4**,  n = 243^1^ |
| **HR+** | 528 (54%) | 0 | 103 (42%) | 133 (55%) | 153 (63%) | 139 (57%) |
| **HER2+** | 244 (25%) | 0 | 35 (14%) | 71 (29%) | 65 (27%) | 73 (30%) |
| **MP2** | 476 (49%) | 0 | 168 (69%) | 128 (52%) | 97 (40%) | 83 (34%) |
| **Immune+** | 446 (46%) | 0 | 140 (57%) | 103 (42%) | 101 (42%) | 102 (42%) |
| **DRD+** | 364 (37%) | 3 | 128 (52%) | 94 (39%) | 71 (29%) | 71 (29%) |
| **PAM50 Subtype** |  | 15 |  |  |  |  |
| LumA | 172 (18%) |  | 16 (6.6%) | 36 (15%) | 57 (24%) | 63 (26%) |
| Basal | 406 (42%) |  | 154 (64%) | 100 (42%) | 82 (34%) | 70 (29%) |
| Her2 | 140 (15%) |  | 27 (11%) | 35 (15%) | 34 (14%) | 44 (18%) |
| LumB | 218 (23%) |  | 42 (17%) | 66 (28%) | 63 (26%) | 47 (20%) |
| Normal | 23 (2.4%) |  | 2 (0.8%) | 2 (0.8%) | 3 (1.3%) | 16 (6.7%) |
| **pCR** | 313 (32%) | 0 | 86 (35%) | 74 (30%) | 79 (33%) | 74 (30%) |
| **Trial Arm** |  | 0 |  |  |  |  |
| Control | 205 (21%) |  | 55 (23%) | 45 (18%) | 53 (22%) | 52 (21%) |
| AMG386 | 133 (14%) |  | 30 (12%) | 37 (15%) | 27 (11%) | 39 (16%) |
| Ganetespib | 93 (9.5%) |  | 28 (11%) | 22 (9.0%) | 27 (11%) | 16 (6.6%) |
| Ganitumab + metformin | 105 (11%) |  | 32 (13%) | 29 (12%) | 26 (11%) | 18 (7.4%) |
| MK2206 | 94 (9.7%) |  | 24 (9.8%) | 17 (7.0%) | 26 (11%) | 27 (11%) |
| Neratinib | 112 (11%) |  | 24 (9.8%) | 32 (13%) | 28 (12%) | 28 (12%) |
| Pembrolizumab | 69 (7.1%) |  | 8 (3.3%) | 11 (4.5%) | 16 (6.6%) | 34 (14%) |
| Pertuzumab | 44 (4.5%) |  | 7 (2.9%) | 19 (7.8%) | 10 (4.1%) | 8 (3.3%) |
| Trastuzumab-emtansine | 52 (5.3%) |  | 7 (2.9%) | 17 (7.0%) | 15 (6.2%) | 13 (5.3%) |
| Carboplatin + ABT888 | 67 (6.9%) |  | 29 (12%) | 15 (6.1%) | 15 (6.2%) | 8 (3.3%) |
| ^1^n (%)  Control: Paclitaxel (+Trastuzumab if HER2+) followed by Anthracyclines | | | | | | |

| **Supplementary Table 3.** Descriptive statistics of IGFBP7 quartiles in relation to clinicopathological characteristics in SCAN-B | | | | | | |
| --- | --- | --- | --- | --- | --- | --- |
|  |  |  | **IGFBP7 Quartiles** | | | |
|  | All patients^1^ | Missing | **Q1**, n = 1,332^1^ | **Q2**, n = 1,332^1^ | **Q3**, n = 1,331^1^ | **Q4**, N = 1,331^1^ |
| **Age at diagnosis, years** |  | 0 |  |  |  |  |
| -40 | 270 (5.1%) |  | 72 (5.4%) | 51 (3.8%) | 85 (6.4%) | 62 (4.7%) |
| 41-50 | 874 (16%) |  | 176 (13%) | 235 (18%) | 260 (20%) | 203 (15%) |
| 51-60 | 1,045 (20%) |  | 233 (17%) | 253 (19%) | 282 (21%) | 277 (21%) |
| 61-70 | 1,661 (31%) |  | 396 (30%) | 426 (32%) | 403 (30%) | 436 (33%) |
| 71-80 | 995 (19%) |  | 279 (21%) | 249 (19%) | 211 (16%) | 256 (19%) |
| 81- | 481 (9.0%) |  | 176 (13%) | 118 (8.9%) | 90 (6.8%) | 97 (7.3%) |
| **Invasive tumor size** (pT2/3/4) | 1,783 (35%) | 166 | 530 (41%) | 421 (32%) | 403 (31%) | 429 (33%) |
| **Lymph node status** (pN1/2/3) | 1,873 (37%) | 213 | 461 (36%) | 475 (37%) | 497 (39%) | 440 (34%) |
| **Histological type** |  | 37 |  |  |  |  |
| Ductal | 4,182 (79%) |  | 1,083 (82%) | 1,085 (82%) | 1,083 (82%) | 931 (71%) |
| Lobular | 732 (14%) |  | 108 (8.2%) | 137 (10%) | 167 (13%) | 320 (24%) |
| Other or Mixed | 375 (7.1%) |  | 131 (9.9%) | 100 (7.6%) | 77 (5.8%) | 67 (5.1%) |
| **Histological grade** |  | 382 |  |  |  |  |
| I | 791 (16%) |  | 130 (11%) | 181 (15%) | 224 (18%) | 256 (21%) |
| II | 2,443 (49%) |  | 512 (42%) | 621 (50%) | 626 (50%) | 684 (55%) |
| III | 1,710 (35%) |  | 588 (48%) | 437 (35%) | 391 (32%) | 294 (24%) |
| **ER+** | 4,497 (85%) | 49 | 1,047 (79%) | 1,138 (87%) | 1,168 (88%) | 1,144 (87%) |
| **PR+** | 3,725 (71%) | 51 | 871 (66%) | 965 (73%) | 959 (72%) | 930 (71%) |
| **HER2+** | 702 (14%) | 149 | 145 (11%) | 192 (15%) | 213 (16%) | 152 (12%) |
| **TNBC** | 525 (10%) | 278 | 208 (17%) | 117 (9.3%) | 91 (7.2%) | 109 (8.6%) |
| **Endocrine therapy** | 4,109 (78%) | 85 | 965 (74%) | 1,046 (79%) | 1,053 (81%) | 1,045 (80%) |
| **Chemotherapy** | 2,243 (43%) | 85 | 616 (47%) | 583 (44%) | 565 (43%) | 479 (36%) |
| **Trastuzumab** | 585 (11%) | 85 | 118 (9.1%) | 165 (13%) | 172 (13%) | 130 (9.9%) |
| **PAM50 Subtype** |  | 0 |  |  |  |  |
| LumA | 2,255 (42%) |  | 365 (27%) | 576 (43%) | 650 (49%) | 664 (50%) |
| Basal | 471 (8.8%) |  | 204 (15%) | 114 (8.6%) | 72 (5.4%) | 81 (6.1%) |
| Her2 | 641 (12%) |  | 131 (9.8%) | 167 (13%) | 193 (15%) | 150 (11%) |
| LumB | 1,275 (24%) |  | 574 (43%) | 391 (29%) | 224 (17%) | 86 (6.5%) |
| Normal | 684 (13%) |  | 58 (4.4%) | 84 (6.3%) | 192 (14%) | 350 (26%) |
| **PAM50 ROR** |  | 292 |  |  |  |  |
| High | 2,361 (47%) |  | 824 (66%) | 674 (54%) | 507 (40%) | 356 (28%) |
| Intermediate | 738 (15%) |  | 205 (16%) | 185 (15%) | 204 (16%) | 144 (11%) |
| Low | 1,935 (38%) |  | 219 (18%) | 399 (32%) | 554 (44%) | 763 (60%) |
| ^1^n (%) | | | | | | |

| **Supplementary Table 4.** *IGFBP7* (in quartiles) gene expression in relation to clinical outcome in SCAN-B | | | | |
| --- | --- | --- | --- | --- |
|  | **Recurrence-**  **free interval** | | **Distant metastasis-**  **free interval** | |
| **Variables** | **HR**^1^ | **95% CI**^1^ | **HR**^1^ | **95% CI**^1^ |
| **Crude** |  |  |  |  |
| **IGFBP7 Quartiles** |  |  |  |  |
| Q1 | Ref. |  | Ref. |  |
| Q2 | 0.95 | 0.75, 1,21 | 0.99 | 0.76, 1,31 |
| Q3 | 0.92 | 0.72, 1.16 | 0.93 | 0.71, 1.23 |
| Q4 | 0.96 | 0.76, 1.22 | 1.00 | 0.77, 1.32 |
| **Multivariable** |  |  |  |  |
| **IGFBP7 Quartiles** |  |  |  |  |
| Q1 | Ref. |  | Ref. |  |
| Q2 | 1.13 | 0.87, 1.47 | 1.26 | 0.93, 1.71 |
| Q3 | 1.20 | 0.91, 1.58 | 1.37 | 1.00, 1.89 |
| Q4 | 1.37 | 1.04, 1.82 | 1.60 | 1.15, 2.22 |
| **Age** (5-year bin) | 1.00 | 0.99, 1.01 | 1.01 | 1.00, 1.02 |
| **Tumor size** (pT2/3/4) | 1.90 | 1.55, 2.32 | 2.26 | 1.78, 2.85 |
| **Lymph node status** (pN1/2/3) | 1.31 | 1.06, 1.62 | 1.57 | 1.24, 2.01 |
| **Grade III** | 1.27 | 0.98, 1.63 | 1.29 | 0.97, 1.73 |
| **ER+** | 1.52 | 0.94, 2.46 | 1.06 | 0.59, 1.91 |
| **PR+** | 0.90 | 0.69, 1.17 | 0.81 | 0.60, 1.09 |
| **HER2+** | 1.95 | 1.27, 2.99 | 2.09 | 1.32, 3.30 |
| **PAM50 ROR High** | 2.20 | 1.61, 3.01 | 2.47 | 1.71, 3.58 |
| **PAM50 subtype** |  |  |  |  |
| LumA | Ref. |  | Ref. |  |
| Basal | 1.53 | 0.91, 2.58 | 1.68 | 0.93, 3.04 |
| Her2 | 1.60 | 1.06, 2.42 | 1.58 | 0.99, 2.53 |
| LumB | 1.21 | 0.87, 1.70 | 1.29 | 0.87, 1.90 |
| Normal | 1.40 | 0.99, 1.97 | 1.47 | 0.97, 2.23 |
| **Chemotherapy** | 0.77 | 0.59, 1.00 | 0.91 | 0.67, 1.24 |
| **Endocrine therapy** | 0.45 | 0.32, 0.63 | 0.75 | 0.46, 1.21 |
| **Trastuzumab** | 0.30 | 0.18, 0.51 | 0.29 | 0.16, 0.51 |

| **Supplementary Table 5.** *IGFBP7* (continuous) gene expression in relation to clinical outcome in SCAN-B | | | | | |
| --- | --- | --- | --- | --- | --- |
|  | **Recurrence-**  **free interval** | | | **Distant metastasis-**  **free interval** | |
| **Variables** | **HR**^1^ | **95% CI**^1^ | **HR**^1^ | | **95% CI**^1^ |
| **Crude** |  |  |  | |  |
| **IGFBP7 Continuous** | 1.01 | 0.88, 1.16 | 1.02 | | 0.87, 1.20 |
| **Multivariable** |  |  |  | |  |
| **IGFBP7 Continuous** | 1.29 | 1.09, 1.53 | 1.41 | | 1.16, 1.73 |
| **Age** (5-year bin) | 1.00 | 0.99, 1.01 | 1.01 | | 1.00, 1.02 |
| **Tumor size** (pT2/3/4) | 1.92 | 1.57, 2.34 | 2.27 | | 1.80, 2.87 |
| **Lymph node status** (pN1/2/3) | 1.30 | 1.05, 1.60 | 1.56 | | 1.23, 1.99 |
| **Grade III** | 1.27 | 0.99, 1.63 | 1.29 | | 0.97, 1.73 |
| **ER+** | 1.52 | 0.94, 2.46 | 1.05 | | 0.58, 1.91 |
| **PR+** | 0.90 | 0.69, 1.18 | 0.81 | | 0.61, 1.09 |
| **HER2+** | 1.92 | 1.25, 2.93 | 2.07 | | 1.31, 3.26 |
| **PAM50 ROR High** | 2.23 | 1.63, 3.05 | 2.52 | | 1.74, 3.64 |
| **PAM50 subtype** |  |  |  | |  |
| LumA | Ref. |  | Ref. | |  |
| Basal | 1.57 | 0.93, 2.65 | 1.72 | | 0.95, 3.11 |
| Her2 | 1.61 | 1.07, 2.43 | 1.59 | | 0.99, 2.55 |
| LumB | 1.24 | 0.89, 1.73 | 1.31 | | 0.89, 1.93 |
| Normal | 1.37 | 0.97, 1.93 | 1.43 | | 0.94, 2.17 |
| **Chemotherapy** | 0.77 | 0.59, 1.00 | 0.91 | | 0.67, 1.24 |
| **Endocrine therapy** | 0.45 | 0.32, 0.63 | 0.75 | | 0.46, 1.21 |
| **Trastuzumab** | 0.31 | 0.18, 0.52 | 0.29 | | 0.17, 0.52 |
