## Supplementary figures and images for "Low *IGFBP7* expression identifies a subset of breast cancers with favorable prognosis and sensitivity to IGF-1 receptor targeting with ganitumab: Data from I-SPY2 and SCAN-B"

### Supplementary Figure 1

**Supplementary Figure 1.**

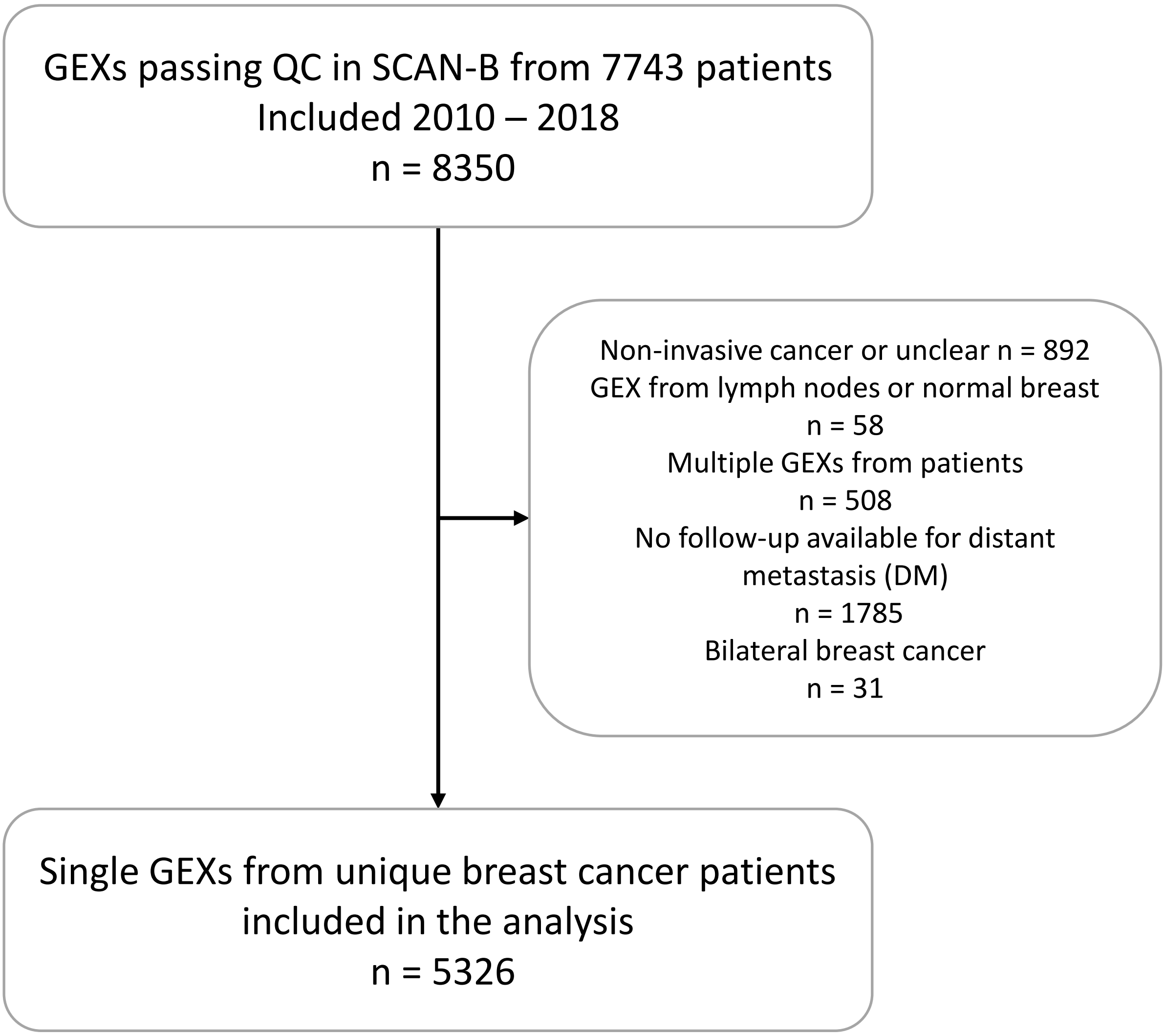

### Supplementary Figure 2

#### Supplementary Figure 2

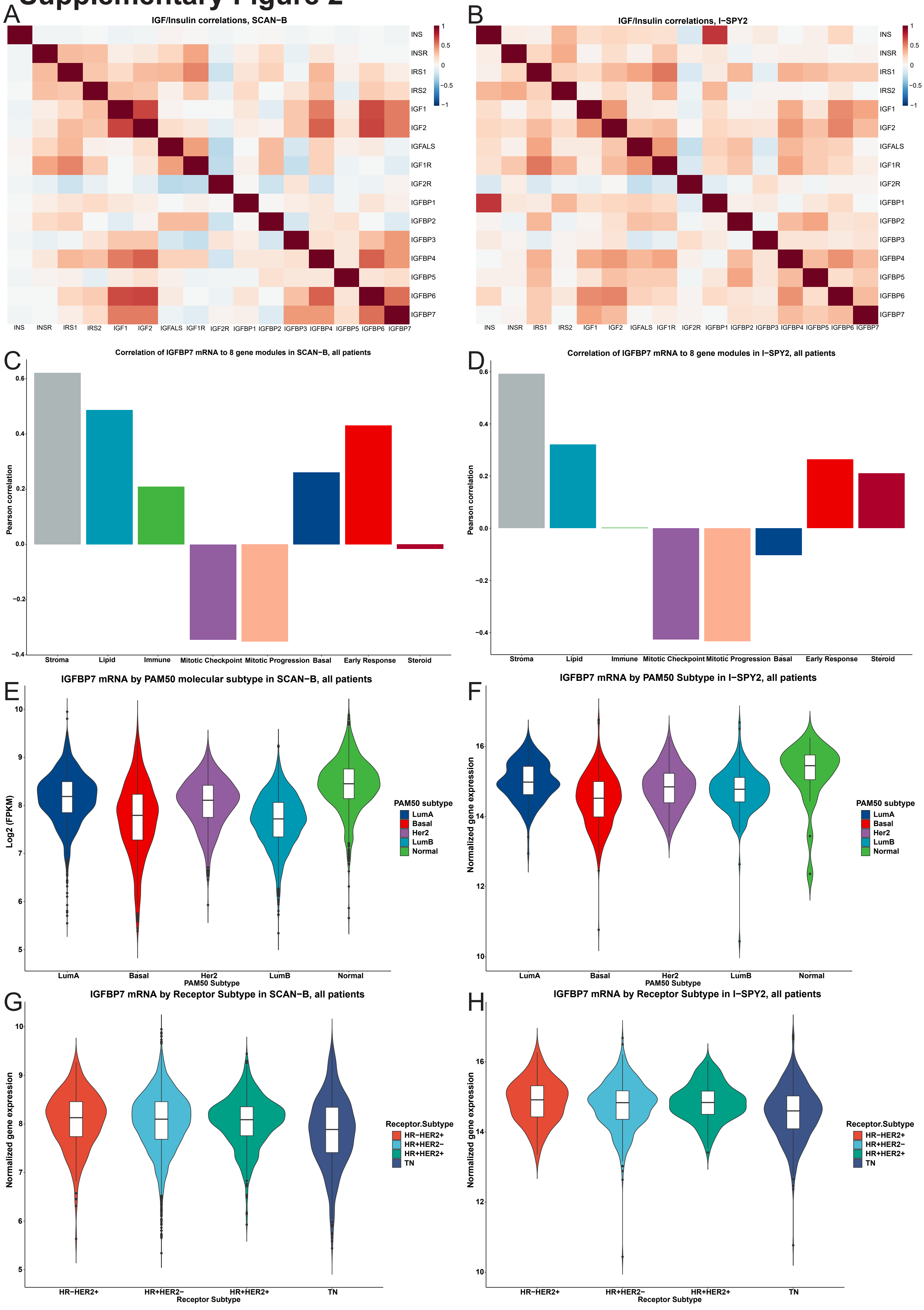
